# Machine Learning-Derived Subtypes of Autism Reveal Distinct Genetic Contributions from Common and Rare Variants

**DOI:** 10.64898/2026.10.19.26365670

**Authors:** Qing Tan, Jingyu Xie, LeeAnne Green Snyder, Runpu Chen, Le Yang, Fei Chen, Brett Trost, Bridget Fernandez, Charleston W. Chiang, Nicholas Mancuso, Chang April Shu

## Abstract

**Background:** Autism spectrum disorder (ASD) is characterized by substantial heterogeneity in core symptom severity and co-occurring conditions, complicating efforts to understand its genetic basis. Most genetic studies rely on binary case-control ASD definitions, which may obscure clinically meaningful variation and distinct genetic contributions across the autism spectrum.

**Methods:** We constructed machine learning (ML)-based categorical ASD subtypes and corresponding continuous ASD subtype scores using 14 key clinical symptom measures with adjustment of sex and age among 6,355 autistic children in the Simons Foundation Powering Autism Research for Knowledge (SPARK) cohort by Discriminative Dimensionality Reduction via learning a Tree (DDRTree). We then replicated and validated these subtypes and scores among 2,717 autistic children in the Simons Simplex Collection (SSC) using a modified k-nearest-neighbor (k-NN) approach. In SPARK and SSC, we further clinically validated the framework by examining associations of subtype classifications and scores with support needs, service utilization and medication use by regression models with adjustment for sex, age, genetic ancestry and socioeconomic factors. We further assessed SNP-based heritability, cell-type-specific heritability enrichment, polygenic scores (PGS) for psychiatric and cognitive traits, as well as rare copy-number variant (CNV) and *de novo* variant (DNV) burdens across the ML-derived ASD subtype scores adjusting for sex, age and genetic ancestry.

**Results:** We identified three clinically distinct ASD subtypes in SPARK varying in the types of supports utilized: a high-support-needs (HSN) subtype characterized by broad developmental and functional impairments and very substantial support needs (OR = 1.63, *P* = 3.71 × 10^-98^); an emotional-behavioral-support-needs (EBSN) subtype marked by severe repetitive behaviors, elevated psychiatric comorbidity; and a low-support-needs (LSN) subtype with milder symptoms and relatively lower overall support needs. The three groups were associated with differential patterns in the specific types of treatments that were most common for them, from higher intensity to lower intensity. Projected ASD subtypes and scores in SSC recapitulated the phenotypic and medication-use patterns observed in SPARK. SNP-based heritability was highest for the EBSN score (h²g = 0.224, SE = 0.051) followed by the HSN (h²g = 0.167, SE = 0.049) and LSN score (h²g = 0.064, SE = 0.044), with suggestive cell-type-specific heritability enrichments in fetal endodermal cells and adult motor neurons for EBSN score and fetal excitatory neurons and astrocytes for HSN score (both *P<*0.05). PGS of ADHD and major depressive disorder were consistently associated with higher EBSN scores in both SPARK and SSC. Higher DNV and rare CNV burden were associated with higher HSN score in SPARK, and higher rare CNV burden was associated with higher EBSN score in SSC (*P<*0.05).

**Conclusions:** Our ML-based transferable ASD subtype framework captured clinically and genetically meaningful dimensions of ASD heterogeneity, warranting further validation in larger cohorts.

## 1. Introduction

Autism spectrum disorder (ASD) is a highly heterogeneous neurodevelopmental condition characterized by differences in social communication and restricted or repetitive behaviors [1,2]. Beyond these core diagnostic features, individuals with ASD vary widely in cognitive ability, language development, behavioral symptoms, adaptive functioning, developmental trajectories and support needs [2–4]. This phenotypic diversity is accompanied by extensive genetic heterogeneity. Both common polygenic variation and rare deleterious variants contribute to ASD risk, but their effects are not expected to be uniform across all individuals with ASD [5–9]. A major challenge in autism research is therefore to move beyond the diagnostic category itself and identify phenotypic dimensions that can clarify how different clinical presentations relate to underlying genetic architecture.

Data-driven approaches have increasingly been used to dissect ASD heterogeneity using multidimensional phenotypic information [10,11]. Studies using clinical phenotypes such as standardized surveys on neurodevelopment and neuroimaging have shown that ASD can be stratified into meaningful subgroups with distinct developmental or biological features [12–14]. More recently, several large-scale studies have demonstrated that phenotype-informed stratification can reveal biologically meaningful heterogeneity within ASD. Warrier et al. identified latent phenotypic dimensions using factor analysis and demonstrated distinct genetic correlates across continuous autism-related traits [8]. Antaki et al. showed that both common polygenic risk and rare deleterious variants contribute to continuous phenotypic variation across ASD [9]. Most recently, Litman et al. applied generative mixture modeling to identify robust phenotypic classes associated with distinct common and rare genetic programs [15]. Together, these studies have established phenotype-driven stratification as a powerful strategy for understanding autism heterogeneity.

However, current subtype frameworks still face several limitations. Some approaches are most effective when comparable phenotypic measures are available across cohorts, limiting their applicability when independent datasets differ in the types or granularity of clinical assessments [8]. They also primarily generate discrete subtype assignments, failing to capture continuous variation between subtype profiles. Furthermore, studies that investigate the genetic architecture of individual symptom dimensions provide valuable biological insights but do not establish an integrated subtype framework that summarizes multidimensional phenotypic heterogeneity into transferable quantitative traits [9]. Thus, a key gap is the lack of a transferable quantitative framework that can define ASD subtype structure in a reference cohort, predict subtype profiles in new individuals and evaluate their phenotypic and genetic validity across datasets.

We hypothesized that continuous subtype scores could provide such a framework. Rather than assigning each individual exclusively to one subtype, continuous subtype scores quantify the extent to which each individual resembles multiple latent subtype profiles. This approach preserves continuous phenotypic variation, captures within-subtype variability and improves power for downstream genetic analyses. Importantly, when learned from a large reference cohort, subtype scores can be predicted in independent individuals, allowing direct cross-cohort comparison and validation within a shared subtype framework.

Here, we applied machine learning (ML) methods to construct ASD subtype classifications and derive continuous ASD subtype scores from clinical symptom measures in the SPARK cohort and characterized these subtypes using service utilization. Next, we projected the ML-derived subtype framework onto autistic individuals in SSC and validated the associated phenotypic patterns. Finally, we investigated the common and rare genetic architectures underlying subtype classifications and continuous subtype scores across both cohorts.

## 2. Methods

### 2.1. Study cohorts and clinical measures

This study focuses on autistic individuals less than 18 years old from the Simons Foundation Powering Autism Research for Knowledge (SPARK, N=6,355) cohort, and the Simons Simplex Collection (SSC, N=2,717). Phenotypic and genetic data from both cohorts were obtained through SFARI Base[16]. SPARK was used as the discovery cohort for developing the ML-derived ASD subtype classifications and continuous scores, while SSC served as an independent validation cohort for evaluating the associated phenotypic and genetic findings.

In SPARK Phenotype Database V10, we included autistic individuals who had complete data for the 14 clinical measures used in the analyses. After removal of one outlier with extremely high values across multiple behavioral measures, the final discovery cohort comprised 6,355 autistic individuals. In SSC Phenotype Database Version 15, autistic probands less than 18 years old with harmonized clinical measures were included. The final validation cohort comprised 2,717 autistic individuals.

### 2.2. Measures on ASD clinical symptoms

In SPARK, 14 measures on ASD core symptoms, co-occurring symptoms and neuropsychiatric conditions were used to construct the ML-derived ASD subtype classifications and continuous subtype scores (**Table S1**).

ASD core symptoms were assessed using the Social Communication Questionnaire-Lifetime (SCQ) summary score [17], and the Repetitive Behavior Scale-Revised (RBS-R) total score[18].

Co-occurring symptoms were assessed by both parent-reported surveys and standardized questionnaires. Cognitive function was measured by three complementary parent-reported measures shown to have relatively good accuracy[19]: (1) cognitive impairment status, indicating whether the individual had ever been diagnosed with intellectual disability or cognitive impairment; (2) cognitive test scores, corresponding to the overall IQ or developmental score from the most recent testing report; and (3) reported categorical cognitive age level (above, at, slightly below or significantly below his/her age). Language level was assessed by parent report using a four-level measure corresponding closely to the ADOS[20] and ADI-R[21]: (1) no words or does not speak; (2) uses single words meaningfully; (3) combines three words into short sentences; and (4) uses longer sentences of his/her own and is able to tell you something that happened. Motor coordination was evaluated using the Developmental Coordination Disorder Questionnaire (DCDQ) total score[22]. Adaptive functioning was evaluated using the Vineland Adaptive Behavior Scales Third Edition - Comprehensive Parent/Caregiver Rating (VABS)[23], including the Adaptive Behavior Composite (ABC), Communication, Daily Living Skills (DLS), and Socialization standard scores, which summarize overall and domain-specific adaptive functions.

Co-occurring neuropsychiatric disorders were defined by parent-reported professional diagnoses of attention-deficit/hyperactivity disorder (ADHD) and mood or anxiety disorders.

In SSC, the same summary scores from standardized questionnaires were used as in SPARK, including summary scores from the SCQ, RBS-R, DCDQ, and VABS Second Edition Comprehensive Interview. Cognitive function was assessed using full-scale IQ (FSIQ) on developmentally appropriate tests. Language ability was assessed using the Autism Diagnostic Interview–Revised (ADI-R) using the following three levels: (1) daily spontaneous use of three-word phrases; (2) functional speech without consistent spontaneous three-word phrase use; (3) fewer than five meaningful words or no functional spoken language. Due to inconsistency in diagnostic practice during the decade of SSC, co-occurring neuropsychiatric disorders were assessed using Child Behavior Checklist (CBCL)[24] T-scores: presence of ADHD, anxious or depressed problems, affective problems, and anxiety problems was defined using the corresponding T-score (T≥70).

### 2.3. Measures on support needs and service utilization

In SPARK, parents rated their child’s daily support needs on a four-level scale: minimal or no support, some support, substantial support and very substantial support. Service utilization was obtained from a parent-reported survey of treatments including speech and language therapy, use of medication, counseling or psychotherapy, and social skills intervention.

In SSC, parent-reported medication history included both current and previous use of antidepressants, antiepileptics, medications for ADHD, sedatives and mood stabilizers or antipsychotics.

### 2.4. ML-derived ASD subtype classifications

ML-derived subtype classifications were constructed in the SPARK discovery cohort using age- and sex-adjusted symptom measures. Autistic individuals were projected into a two-dimensional space with a latent tree structure using the discriminative dimensionality reduction with trees (DDRTree) methods[25] as implemented in Monocle2[26]. Default hyperparameters recommended by DDRTree[26] were retained for parameters controlling optimization and regularization (e.g., **sigma**, **param.gamma**, and **tol**), while the maximum number of iterations (**maxIter = 30**) was chosen to ensure convergence without unnecessary computation. The number of graph centers (**center = 64**), which controls the complexity of the learned tree structure, was selected empirically after evaluating multiple parameter settings. Smaller values tended to oversimplify the tree structure by merging distinct phenotypic patterns, whereas larger values generated increasingly complex branching structures that reduced interpretability. The final parameter setting produced a stable three-branch tree structure with balanced sample sizes across branches. Autistic individuals with similar profiles are grouped based on whether they are assigned to the same underlying branches of the tree structure.

To statistically evaluate the internal validity of ML-derived ASD subtypes, three complementary clustering metrics were calculated: the silhouette score[27], Davies–Bouldin index (DBI)[28], and Calinski–Harabasz index (CHI) [29]. The silhouette score evaluates both within-cluster cohesion and between-cluster separation by comparing the similarity of each individual to its assigned cluster relative to neighboring clusters, with higher values indicating better-defined clusters[27]. The DBI measures the average similarity between each cluster and its most similar neighboring cluster based on within-cluster dispersion and between-cluster distance, with lower values indicating better cluster separation and less overlap[28]. The CH index quantifies the ratio of between-cluster dispersion to within-cluster dispersion, with higher values indicating better overall clustering performance [29]. These metrics were used to compare the internal validity of three- and four-branch subtype clusters by DDRTree and to compare their performances with k-means clustering based on the same 14 standardized clinical measures.

To clinically evaluate the ML-derived ASD subtypes, we assessed the clinical relevance of the patterns across subtypes and the support needs as described in Methods 2.7 and 2.8, and refined the parameters in DDRTree to ensure the overall pattern and the latent tree structures to align with clinical observations.

### 2.5. ML-derived ASD subtype scores

In addition to ASD subtype classifications, we derived continuous ASD subtype scores using the trajectory inference framework implemented in Monocle 2[26]. Autistic individuals were ordered based on symptom severity and comorbidities along branches of the tree structure using DDRTree.

Continuous subtype scores were derived by applying the Monocle 2 trajectory-ordering procedure (a.k.a, pseudotime) separately to each subtype branch [26]. For each subtype, the terminal node of the corresponding branch was designated as the trajectory root, and pseudotime was recalculated for all autistic individuals. Pseudotime was defined as the distance from the selected root along the inferred tree structure, rather than the Euclidean distance in the two-dimensional space by DDRTree. The resulting pseudotime values were normalized to range from 0 to 1, yielding a continuous subtype score for each individual. For each subtype, these scores quantify an individual’s relative position along the corresponding branch of the inferred latent tree, with larger values indicating greater proximity to that subtype.

### 2.6. Projecting autistic individuals in SSC onto the ML-derived ASD subtype framework

To project autistic individuals in SSC onto the ML-derived ASD subtype framework developed in SPARK, we developed a modified k-nearest-neighbor (kNN) mapping approach to assign each individual to a subtype and compute the corresponding subtype score. We provided a summary of the methods below with further details available in the **Supplementary Notes and Supplementary Figure 1**.

First, missing clinical measures in SSC were imputed using MissForest [30], which was previously shown to achieve good imputation accuracy for summary scores in SPARK [31]. Specifically, SCQ summary scores were imputed for 823 individuals (28.7%) and DCDQ total scores for 471 individuals (16.4%). The top 500 clinical variables most strongly correlated with both SCQ and DCDQ and with <20% missingness were used as predictors. To evaluate imputation performance in SSC, we used a subset of individuals with complete SCQ and DCDQ data, randomly masked observed values according to their corresponding missing rates, and compared the imputed and observed values using mean squared error and Pearson correlation as described previously [31].

Next, symptom measures in SSC were standardized using the mean and standard deviation of the corresponding measures in SPARK. Measures of the same symptom domain using different instruments were harmonized. Specifically, measures of language ability comprised four levels in SPARK and three levels in SSC. There was substantial missingness of CBCL scores in SPARK, and parental-reported history of ADHD and mood disorders were used. Language measures from the two cohorts were defined as ordinal values reflecting increasing language impairment and normalized to a 0-1 scale. CBCL T-scores for attention, ADHD, and mood or anxiety symptoms in SSC were normalized to a 0-1 scale to enable cross-cohort projection with the corresponding binary medical history indicators in SPARK.

For each autistic individual in SSC, the three most similar autistic individuals in SPARK were identified by Euclidean distance based on the shared standardized clinical measures. Within each subtype branch, root-to-node distances of neighboring SPARK participants were averaged using proximity-based weights so that more phenotypically similar autistic individuals in SPARK contributed more strongly than more distant participants. Because the three subtype branches differed in their geometric scales, weighted distances were normalized using subtype-specific branch-scale factors and transformed into subtype-specific similarity scores. These similarity scores jointly incorporated local neighborhood composition, phenotypic similarity, and the geometry of the reference DDRTree. The predicted subtype assignment for each SSC participant was defined as the subtype with the highest similarity score. Continuous subtype scores were estimated as similarity-weighted averages of the corresponding subtype scores from neighboring SPARK participants. Further details are available in the **Supplementary Notes**.

The reliability of the projected subtype assignments was evaluated for neighborhood sizes (*k*) ranging from 2 to 5 using five stratified bootstrap replicates in SPARK. Reliability was defined according to the consistency of subtype assignments across bootstrap replicates: complete agreement across all five replicates was classified as high reliability, agreement in four replicates as moderate reliability, and agreement in three or fewer replicates as low reliability. The variability of the projected subtype scores was evaluated by the standard deviation (SD) of subtype scores across five bootstrap replicates. For each neighborhood size, the mean of these individual bootstrap SDs across all SSC participants was computed separately for the HSN, EBSN, and LSN scores, with lower mean SDs indicating greater reliability of the projected subtype scores. Although mean bootstrap SDs decreased modestly as increasing neighborhood size, differences across neighborhood sizes were small, indicating that the projected subtype scores were robust to neighborhood size selection (**Table S3a**). We selected k = 3 as the final neighborhood size for kNN-based projection since it showed the highest combined proportion of individuals with high or moderate reliability and low SDs of the projected subtype scores across five bootstrap replicates(**Table S3b**).

### 2.7. Association of support needs and service utilization with ML-derived ASD subtypes

Associations between subtype class/scores and each support or intervention outcome were evaluated using logistic regression, adjusting for age at registration, sex, the first 10 genetic ancestry principal components (PCs), both parents’ education and household income. Pairwise subtype comparisons were performed using Subtype 3 (low-support-needs subtype) as the reference group. Odds ratios (ORs), 95% confidence intervals (CIs), and two-sided *P* values were reported.

### 2.8. Association of DSM-IV diagnoses with ML-derived ASD subtypes

Parent-reported DSM-IV autism diagnoses included Autistic Disorder, Asperger’s Disorder, and Pervasive Developmental Disorder–Not Otherwise Specified (PDD-NOS). Associations between DSM-IV diagnostic categories and ML-derived ASD subtype assignments were evaluated using Pearson’s χ² tests.

### 2.9. Quality control and imputation for genotyping data in SPARK

Genotyping data from 5,309 autistic individuals from the three batches (Array v1, Array v2, and GxS) were obtained from SPARK iWES v3 in SFARI Base. These batches were preprocessed separately using the quality-control and imputation pipeline described as follows. Samples with call rates < 90% and sex discordance were removed, and variants with genotype missingness > 5% were removed. For autistic individuals with available parental genotype data, reported familial relationships were validated using KING v2.2.7 [32]. Individuals and families with excessive Mendelian errors (Z score > 3) and variants with Mendelian error rates greater than 10^-4^ were removed using PLINK v1.9 [33]. Genetic ancestry was inferred using Neural Admixture v1.5.1 [34], and genotype data were imputed separately for each inferred ancestry group within each batch using the TOPMed r3 reference panel by TOPMed Imputation Server [35].

All imputed genotyping data were subsequently merged and a combined R^2^ indicating overall imputation quality was computed using IMMerge v0.0.4 [36]. Post-imputation quality control retained variants with combined R^2^ ≥ 0.6. For variants mapping to the same genomic position, the variant with the highest combined R^2^ was retained, and variants with overall genotype missingness > 5% were excluded. Ancestry PCs among variants with MAF<0.05 were computed by PLINK 1.9 [33].

### 2.10. Quality control for whole genome sequencing data in SSC

Whole-genome sequencing (WGS) data from 2,385 autistic individuals in the Simons Simplex Collection (SSC) were obtained from SFARI Base. Common variants with genotype missingness > 5% or minor allele frequency (MAF) < 1% were excluded using PLINK 1.9 (version, ref). Principal component analysis was performed using autosomal variants after linkage disequilibrium (LD) pruning to characterize population structure.

### 2.11. Copy number variant burden in SPARK and SSC

Copy number variant (CNV) analyses for the SPARK cohort were restricted to autosomal rare CNVs called from whole-exome sequencing (WES) data via XHMM by the SFARI team [37, 38]. CNVs were retained that spanned at least three exons, measured at least 1 kb, and occurred at a frequency below 1% in the source cohort. For each participant, the numbers of total CNVs, deletions, and duplications were calculated and used as sample-level CNV burden measures.

For the SSC cohort, CNVs were obtained from whole-genome sequencing (WGS) data and filtered at the individual CNV-record level using the quality-control criteria defined in the original SSC annotation files and the published WGS CNV framework [39]. CNVs were retained when they passed the original call filter, were detected by both CNVnator[40] and ERDS [41], with the same CNV type and at least 50% reciprocal overlap, and overlapped difficult genomic regions by less than 70%. Analyses were further restricted to rare CNVs with a frequency below 1% in the source cohort and a genomic length of at least 1 kb.

CNV burden was tested in relation to ML-derived ASD subtypes using two complementary approaches. First, subtype classification was modeled as a function of total CNV burden, deletion burden, or duplication burden using multinomial logistic regression models. Second, subtype scores were modeled as a function of total CNV burden, deletion burden, or duplication burden using linear logistic regression models. All models were adjusted for sex, age at registration, and the first ten ancestry PCs.

### 2.12. *De novo* variant burden in SPARK and SSC

High-confidence *De novo* variants (DNVs) in SPARK and SSC were obtained from Zhou et al. (2022) [42] following standardized quality-control, annotation, and variant-filtering procedures. For each autistic individual, we quantified the total number of DNVs and the number of loss-of-function (LoF) DNVs. Associations between DNV burden and subtype assignments were evaluated using multinomial logistic regression models, adjusting for age at registration, sex and the first 10 genetic ancestry PCs, whereas associations with logit-transformed continuous subtype scores were evaluated using linear regression with the same covariate adjustments.

### 2.13. SNP heritability

SNP-based heritability of the ML-derived subtype scores was estimated in SPARK using common variants after quality control as previously described using BOLT-REML [43]. In SPARK, 3,448,423 variants with MAF > 1% and genotype missingness < 5% were retained for heritability estimation.

Kinship was estimated using KING [32], and 266 and 1 related individuals were removed in SPARK and SSC, respectively. The resulting dataset was used to estimate SNP-based heritability for each subtype score using BOLT-REML. All models included sex, age, and the first ten ancestry PCs as fixed-effect covariates.

### 2.14. Cell type heritability enrichment

Cell-type heritability enrichment analyses were performed using CT-FM[44]. GWAS summary statistics for the logit-transformed subtype scores were generated using BOLT-LMM [43], with adjustment for sex, age at registration, and the first ten ancestry PCs. These summary statistics were then used to estimate partitioned SNP heritability across cell-type-specific cis-regulatory annotations using stratified linkage disequilibrium score regression (S-LDSC) [45].

To improve biological interpretability and reduce redundancy among highly related annotations, the original 927 candidate cis-regulatory element (cCRE) annotations were manually curated and merged into canonical cell-type categories according to developmental stage and cell-type name. For annotations derived from the Zhang et al. [46], numerical suffixes denote distinct chromatin accessibility clusters within the same annotated cell type. Cell-type enrichment was evaluated using the S-LDSC regression coefficient Z score and the enrichment *P* value. Cell type heritability enrichment with on-negative coefficient Z score and enrichment *P* < 0.05 were reported for HSN and EBSN subtype scores.

### 2.15. Polygenic scores

Polygenic scores (PGSs)[47] were calculated for seven psychiatric and cognitive traits relevant to ASD, including ASD, attention-deficit/hyperactivity disorder (ADHD), schizophrenia (SCZ), bipolar disorder (BD), major depressive disorder (MDD), anxiety disorders (ANX), and cognitive function. GWAS summary statistics for ASD, ADHD, SCZ, BD, MDD, and ANX were obtained from the Psychiatric Genomics Consortium (PGC) [48–53], whereas GWAS summary statistics for cognitive function were obtained from Davies et al. [54]. PGSs were generated using PGS-CSx [55] from the imputed genotype dataset and were standardized to Z-scores prior to downstream analyses.

Associations between PGSs and subtype assignments were evaluated using multinomial logistic regression, whereas associations between PGSs and logit-transformed continuous subtype scores were evaluated using linear regression. Separate association analyses were performed for each PGS. All models included sex, age, and the first ten ancestry PCs as covariates.

## 3. Results

### 3.1. Overview

**Figure 1** provides an overview of the analytical pipeline used in this study. Using DDRTree[25], ML-derived ASD subtype classifications and continuous subtype scores were constructed using clinical measures from 8 symptom domains in the SPARK cohort as the discovery set (N=6,355). Subsequently, we projected the ML-derived ASD subtypes to the SSC cohort (N=2,717) as the validation set. We then assessed the common and rare genetic burden across ML-derived ASD subtypes, including SNP heritability, cell-type heritability enrichment, polygenic risk scores, rare copy number variants, and *de novo* mutations.

**Figure 1.**
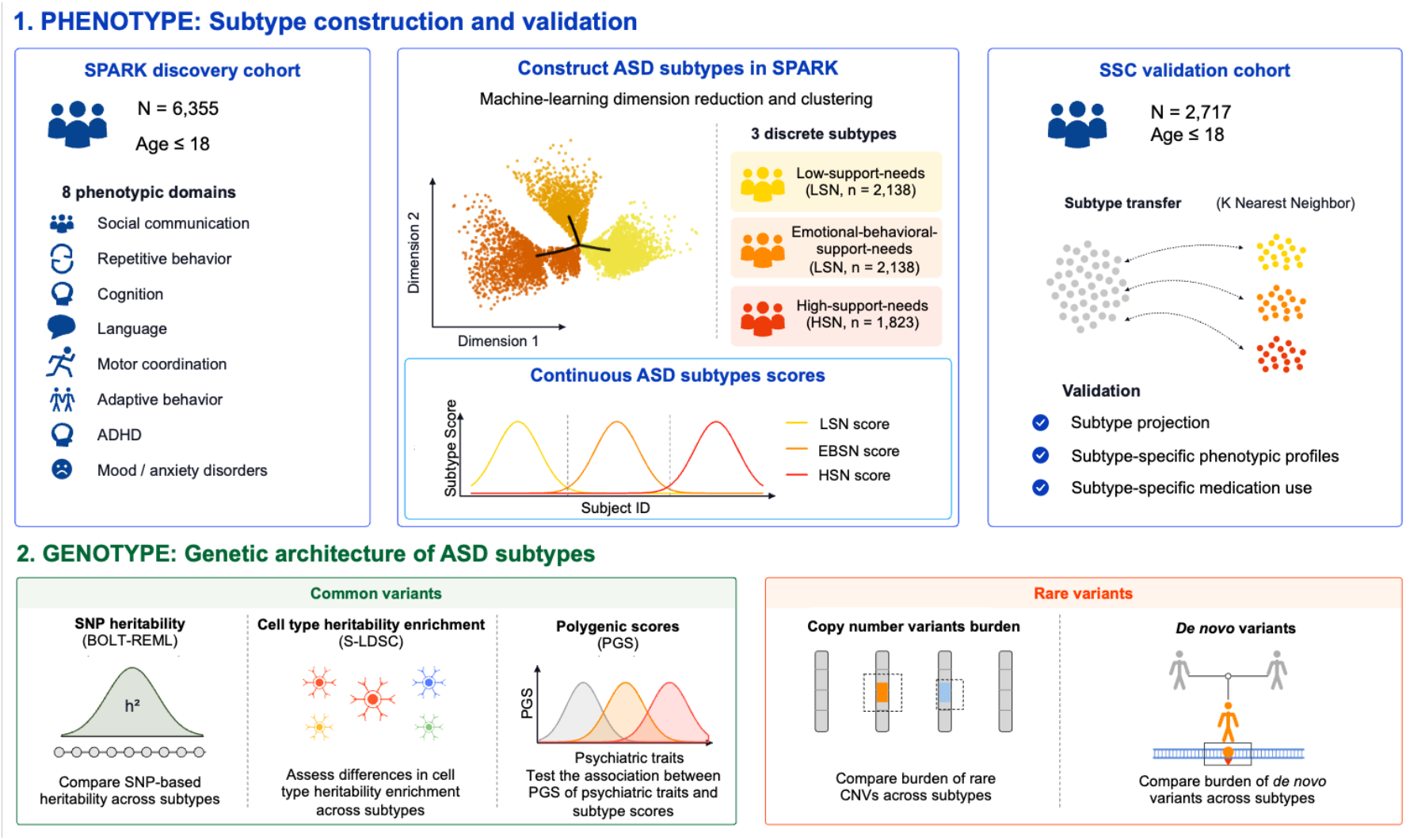
Overview of the study design and analytical framework. First, machine learning (ML)-derived ASD subtypes were constructed among individuals with ASD aged ≤ 18 years in the SPARK study (N = 6,355) using clinical measures in 8 ASD core symptoms, co-occurring symptoms and disorders. DDRTree [25] was applied to derive three ASD subtype classifications and continuous subtype scores. Second, autistic individuals aged ≤ 18 evaluated service data in SSC cohort (N = 2,717) were projected to ML-derived ASD subtypes in SPARK using a k-NN approach as validation. Third, common and rare genetic burden analyses were performed to characterize subtype-specific genetic architectures, including SNP-based heritability, cell-type heritability enrichment, polygenic risk scores, rare copy number variant burden and de novo mutation burden analyses.

### 3.2. ML-derived ASD subtypes

Among 6,355 autistic individuals aged ≤ 18 years in SPARK, we selected 14 clinical measures with complete data on ASD core symptoms, co-occurring symptoms and disorders based on SCQ, RBS-R, DCDQ, VABS, as well as parent-reported symptoms and medical history (**Table S1**). We standardized these measures by age and sex, and constructed a two-dimensional DDRTree plot, where autistic individuals were ordered based on symptom severity and comorbidities along branches of the latent tree structure indicating three distinct ASD subtypes using DDRTree (**Figure 2A**). The ML-derived ASD subtypes demonstrated better internal clustering validity compared with k-means clustering (**Table S2)**. Sensitivity analysis of an alternative four-subtype solution showed reduced cluster cohesion and separation compared with the three-subtype solution (**Table S2**).

**Figure 2.**
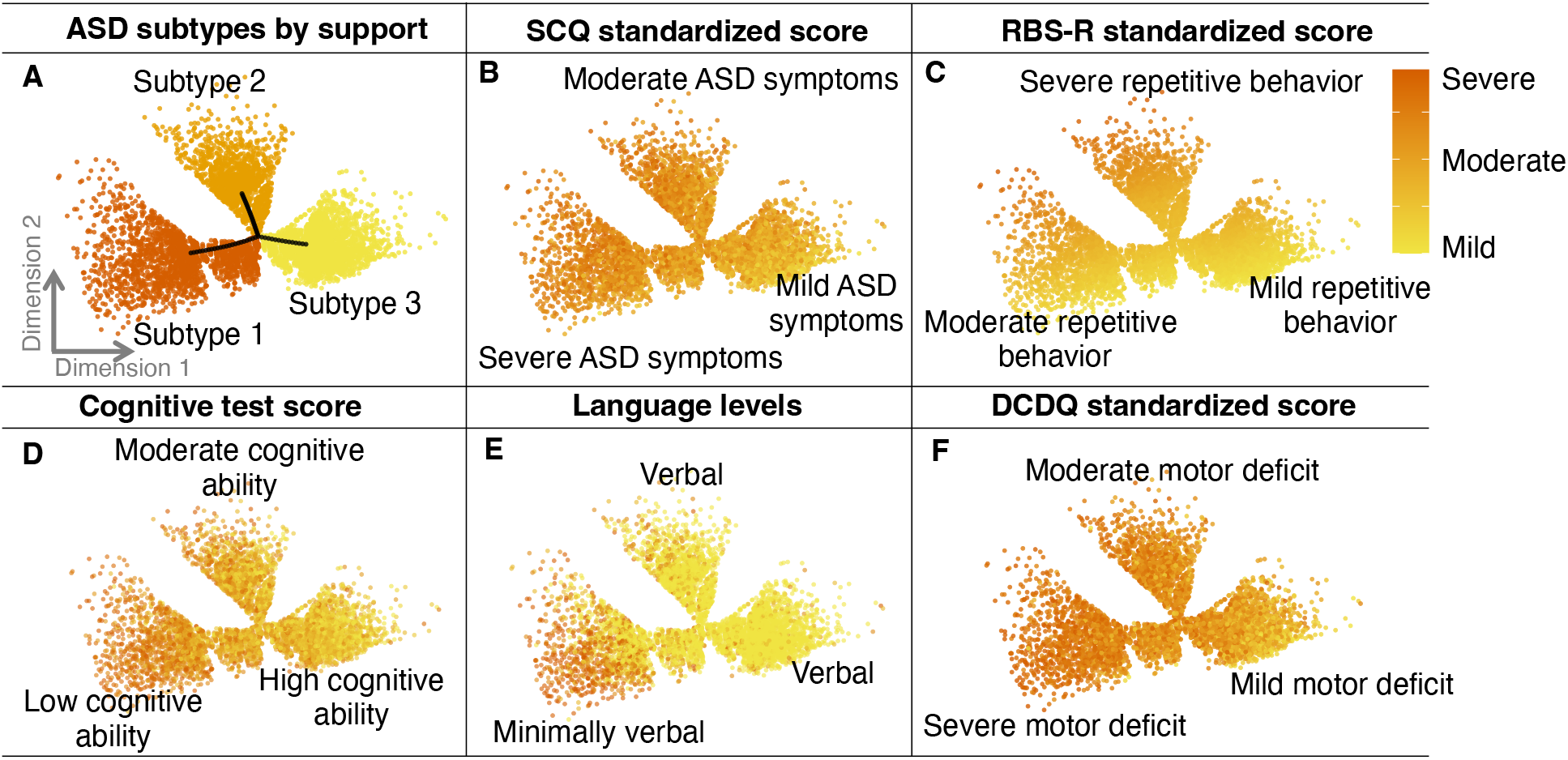
Clinical profiles underlying ML-derived ASD subtypes. Panel A shows the distribution of the three ASD subtypes, whereas B-F displays patterns of ASD symptoms (SCQ), restricted and repetitive behaviors (RBS-R), cognitive function, language ability, and motor coordination (DCDQ) for each subtype. Across multiple clinical domains, symptom burden and functional impairment changed gradually across the subtype landscape rather than abruptly between groups. These findings suggest that the identified support subtypes represent major regions within a broader continuum of ASD related phenotypic variation.

ML-derived ASD subtypes showed distinct patterns of symptom severity. **Subtype 1** (red) was characterized by broad developmental and functional difficulties, including higher impact of ASD symptoms and impairments in cognitive and language abilities (**Figures 2 B-F, Table 2**). **Subtype 2** (orange) showed the highest impairment in repetitive behaviors and elevated co-occurring psychiatric disorders including ADHD, mood and anxiety disorders (**Figures 2B-2F**, **Table 2**).

**Subtype 3** (yellow) showed comparatively milder symptom severity across these domains and a lower burden of co-occurring psychiatric disorders (**Figures 2B-2F**, **Table 2**).

### 3.3. Continuous ASD Subtype Scores

In addition to categorical subtype classifications, we constructed continuous ASD subtype scores on a scale of 0 to 1 for each subtype. Scores were calculated from participants’ latent positions in the tree structure relative to the most severely affected individual within each subtype (**Figure 3**). A score closer to 1 indicates greater similarity to the corresponding subtype profile, whereas a score closer to 0 indicates less similarity. We examined the associations between these scores and the core phenotypic features used to derive the subtypes and observed patterns consistent with the categorical classifications. Subtype 1 scores (**Figure 3** panel A-C, **Table 3**) were positively associated with ASD symptoms (r = 0.37, *P* < 2 × 10^-16^), and negatively associated with cognitive test scores (r = −0.49, *P* < 2 × 10^-16^) and ADHD incidence (Δ= −0.02, *P* < 5 × 10^-6^), and showed negligible association with repetitive behavior (r = 0.10, *P* < 2 × 10^-16^). Conversely, Subtype 2 scores (**Figure 3** panel D-F, **Table 3**) were strongly positively associated with repetitive behavior scores (r = 0.48, *P* < 2 × 10^-16^) and ADHD incidence (Δ = 0.05, *P* < 2 × 10^-16^) but less so with ASD symptoms (r = 0.19, *P* < 2 × 10^-16^), showed minimal correlation with cognitive test scores (r= 0.099, *P* = 3 × 10^-15^) and were associated with higher ADHD incidence (Δ = 0.05, *P* < 2 × 10^-16^_)._

**Figure 3.**
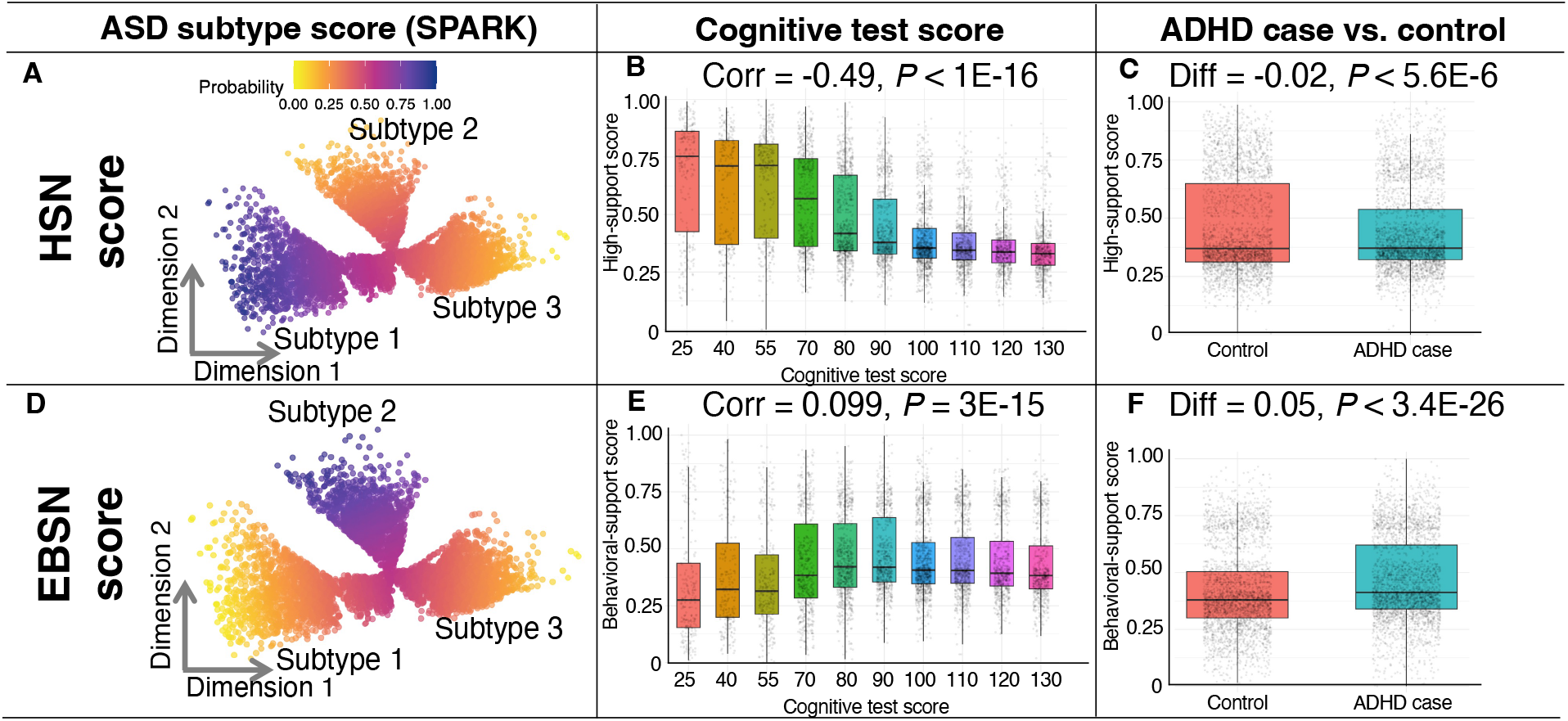
ML-derived ASD subtype scores and their associations with cognitive test scores and ADHD incidence. The panel A-C showed high-support-needs (HSN) scores and the panel D-F showed emotional-behavioral-support-needs (EBSN) scores (range: 0-1). HSN scores were negatively associated with cognitive test score and slightly lower ADHD incidence, whereas EBSN scores showed near-zero correlation with cognitive test score and higher ADHD incidence.

### 3.4. Clinical Validation of ML-derived ASD Subtypes and Scores

Several real-world clinical indicators, which were not included in the construction of the ML-derived ASD subtype assignment or subtype scores, were used to validate the ML-derived ASD subtypes and their scores. ASD subtypes showed a differential pattern of association with the clinical DSM-IV diagnostic subtypes. Of individuals in SPARK with a previous DSM-IV diagnosis of Asperger’s Disorder, which captures individuals with higher verbal and cognitive ability, 59.8% were assigned to Subtype 3, whereas only 10.8% and 29.4% were assigned to Subtypes 1 and 2, respectively (*P* = 5.28 × 10^-30^).

Support needs and service utilization exhibited distinct subtype-specific patterns (**Figure 4**, **Table 4**). Autistic individuals in Subtype 1 exhibited the highest reported support needs, with 44.0% requiring substantial support and 24.9% requiring very substantial support; only 3.9% required minimal or no support (**Figure 4**). Autistic individuals in Subtype 2 showed intermediate support needs, with most individuals requiring some (39.6%) or substantial (42.8%) support (**Figure 4**). Subtype 3 had the lowest support needs compared with other subtypes, with the majority of the Subtype 3 group requiring some (58.3%) or minimal to no (20.1%) support (**Figure 4**). In terms of the ASD subtype scores, similar patterns of association with support needs were found using logistic regression, even after adjusting for age, sex, the first 10 genetic ancestry PCs, family income, and parental education (**Table 4**). Specifically, a 0.1-unit increase in Subtype 1 score was associated with 1.63-fold higher odds of requiring very substantial support (OR = 1.63, *P* = 3.71 × 10^-98^), whereas a 0.1-unit increase in Subtype 3 score was associated with 1.90-fold higher odds of minimal or no support (OR = 1.90, *P* = 5.25 × 10^-75^).

**Figure 4.**
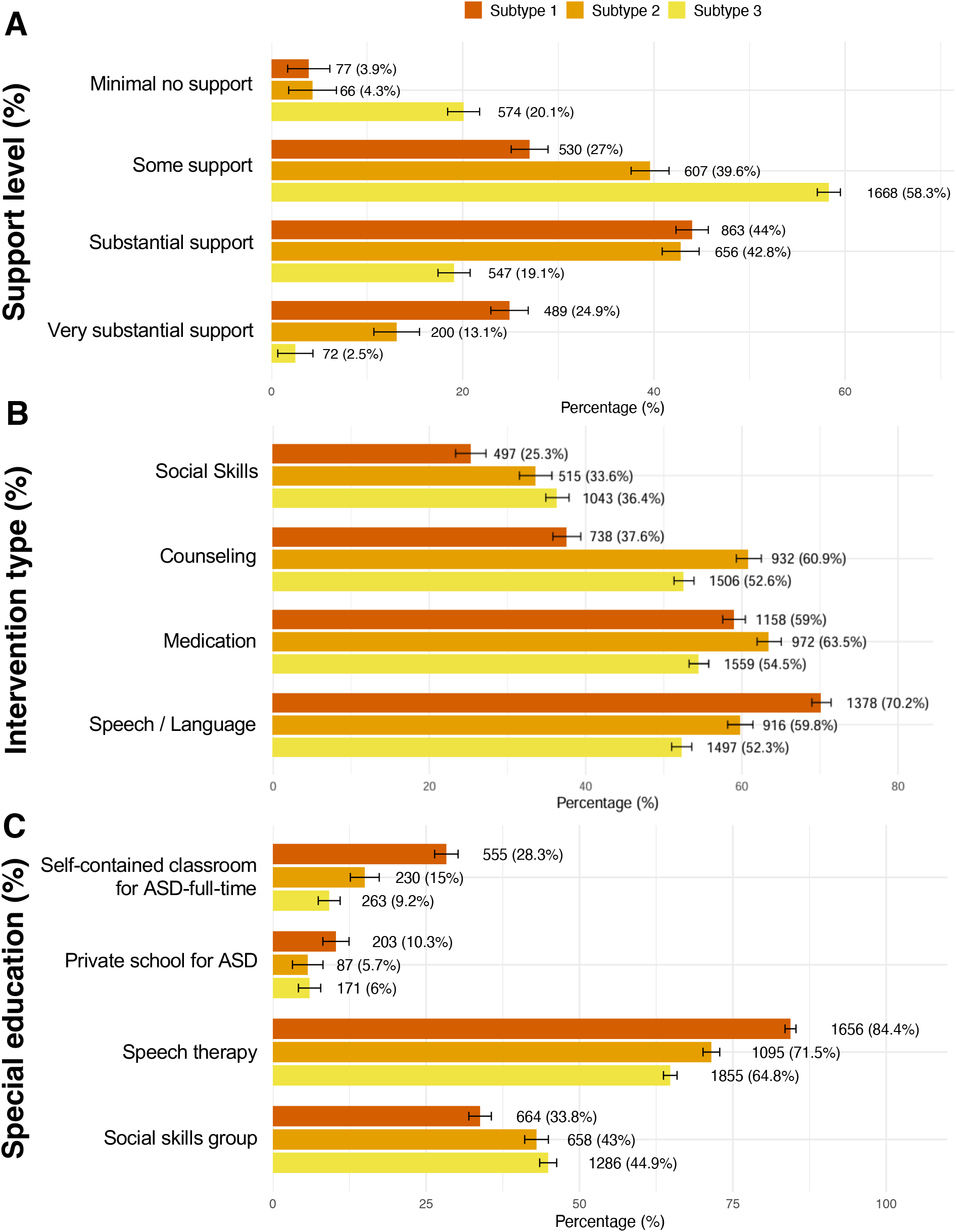
Support needs and use of private interventions across ML-derived ASD subtypes. Panel A shows the distribution of support needs ranging from minimal or no support to very substantial support across ML-derived ASD subtypes. Panel B shows the percentage of subjects in each subtype receiving different types of private interventions, including speech or language therapy, medication, counseling, and social skills support. Panel C shows the percentage of subjects in each subtype receiving selected special education services, including placement in a self-contained classroom for ASD on a full-time basis, attendance at a private school for ASD, speech therapy, and participation in a social skills group. Bars represent percentages within each subtype, with numbers indicating the corresponding sample counts. Error bars indicate 95% confidence intervals.

Patterns of private intervention use also differed across subtypes (**Figure 4**, **Table 4**). Autistic individuals in Subtype 1 were the most likely to receive speech-language therapy (70.2%), and were also more likely to receive medication than those in Subtype 3. Subtype 2 had the highest rates of counseling (60.9%) and medication use (63.5%), amongst all subtypes (**Figure 4**). The associations between each type of private intervention and ASD subtype scores were also supported by logistic regression after adjustment for age, sex, the first 10 ancestry PCs, family income, and parental education (**Table 4**). A 0.1-unit increase in Subtype 1 score increases the odds of receiving speech-language therapy by 21% (OR = 1.21, *P* = 1.71 × 10^-30^), whereas a 0.1-unit increase in Subtype 2 score increases the odds of receiving counseling as compared to other types of interventions (OR = 1.23, *P* = 1.21 × 10^-30^) and medication use (OR = 1.10, *P* = 6.48 × 10^-8^). The association between social skill support and 0.1-unit increase in Subtype 3 score was attenuated but remained strong after adjusting for demographic and socioeconomic factors (OR= 1.05, *P* = 2.63 × 10^-3^), whereas 0.1-unit increase in Subtype 2 score showed the strongest association with social skill support (OR= 1.11, *P*=1.27 × 10^-8^).

Additionally, special education service use differed across the ML-derived ASD subtypes. After adjusting for demographic and socioeconomic factors, a 0.1-unit increase in Subtype 1 score was consistently associated with higher odds of placement in a full-time self-contained classroom for ASD or other diagnosis, a private school for ASD, or with a one-on-one classroom aide (all *P* < 1.03 × 10^-14^, **Table 4**). Consistent with our findings in private interventions, a 0.1-unit increase in Subtype 1 score was associated with greater use of speech therapy (OR = 1.30, *P* = 6.16 × 10^-39^, **Table 4**), whereas a 0.1-unit increase in Subtype 2 score and Subtype 3 score were associated with participation in social skill groups (Subtype 2: OR = 1.13, *P* = 5.30 × 10^-13^; Subtype 3: OR= 1.05, *P* = 2.77 × 10^-3^, **Table 4**). Higher 0.1-unit increase in Subtype 1 score was also associated with receiving occupational and physical therapy (all *P* < 8.71 × 10^-40^).

Taken together, these findings indicate that the three ML-derived ASD subtypes capture clinically meaningful differences in symptom profiles, psychiatric comorbidity, and support needs among individuals with ASD. Accordingly, we designate Subtype 1 as the high-support-needs (HSN) subtype, reflecting broad developmental impairments and the greatest overall service utilization; Subtype 2 as the emotional-behavioral-support-needs (EBSN) subtype, characterized by the highest burden of co-occurring psychiatric symptoms, and the greatest need for behavioral interventions; and Subtype 3 as the low-support-needs (LSN) subtype, representing individuals with comparatively milder symptoms, lower overall support needs but greatest needs in social skills support. These designations are used throughout the subsequent analyses to facilitate clinical interpretation of the ML-derived subtypes.

### 3.5. ML-derived ASD subtypes generalize to an independent ASD cohort

Autistic individuals in SPARK and SSC had similar mean age at registration, with female participants comprising 24.0% of SPARK and 13.5% of SSC (**Table 1**). SPARK also included slightly higher proportions of African Americans and multiracial participants than SSC (**Table 1**). The two cohorts showed broadly comparable levels of social communication impairment (SCQ: Δ = 1.37, *P* = 5.35 × 10^-20^), motor coordination difficulties (DCDQ: Δ = −0.61, *P* = 2.93 × 10^-3^) and adaptive functioning (VABS ABC: Δ = −0.03, *P* = 5.21 × 10^-2^), whereas repetitive behaviors, co-occurring ADHD and mood disorders were more severe among SPARK participants, as reflected by significantly higher RBS-R total scores (Δ = 8.59, *P* = 3.83 ×10^-77^; **Table 1**), CBCL T-scores for ADHD (Δ = 2.78, *P* = 8.88 × 10^-37^) and anxiety or depression (Δ = 5.23, *P* = 2.42 × 10^-90^; **Table 1**). These differences are consistent with the ascertainment strategies of the two cohorts, as SSC comprises only simplex families and excludes probands with low mental age, abnormality in brain and birth defects [56], whereas SPARK includes both simplex and multiplex families and autistic individuals with severe impairments.

We projected the ML-derived ASD subtype classifications and scores in SPARK into SSC using a k-nearest-neighbor (kNN) approach (**Methods 2.6**). Only 18.9% autistic individuals in SSC were classified as the EBSN subtype compared to 24.1% in SPARK. This was consistent with the cross-cohort differences that autistic individuals in SSC had lower levels of repetitive behaviors and co-occurring ADHD and mood disorders than SPARK.

We assessed the reliability of subtype projections in SSC by bootstrapping the SPARK reference subjects used in the projection (**Methods 2.6**). Overall, 78% of autistic individuals in SSC had concordant subtype assignments across all five bootstrap replicates, 12% across four replicates and 10% across three or fewer replicates (**Table S3b**). Projected scores for all three subtypes also showed low variability across bootstrap replicates, with relatively small standard deviations (**Table S3a**). These findings were consistent across kNN neighborhood sizes ranging from k = 2 to k = 5 in the projection.

The projected ASD subtypes in SSC recapitulated the subtype-specific phenotypic profiles observed in SPARK(**Figure 5**, **Table 2**). The SCQ summary scores among autistic individuals in HSN and EBSN subtypes were significantly higher than those in LSN subtype (*P* < 1 × 10^-100^), whereas the RBS-R summary scores were highest among those in EBSN subtype (*P* < 1 × 10^-100^, **Table 2**). Across both cohorts, autistic individuals in the HSN subtype consistently exhibited the greatest impairment in language ability, cognitive performance, motor coordination, and adaptive functioning, while those in the LSN subtype showed the least impairment (**Table 2**). The EBSN subtype was characterized by the highest severity of repetitive behaviors and co-occurring disorders including ADHD and mood or anxiety disorders in both SPARK and SSC (**Table 2**).

**Figure 5.**
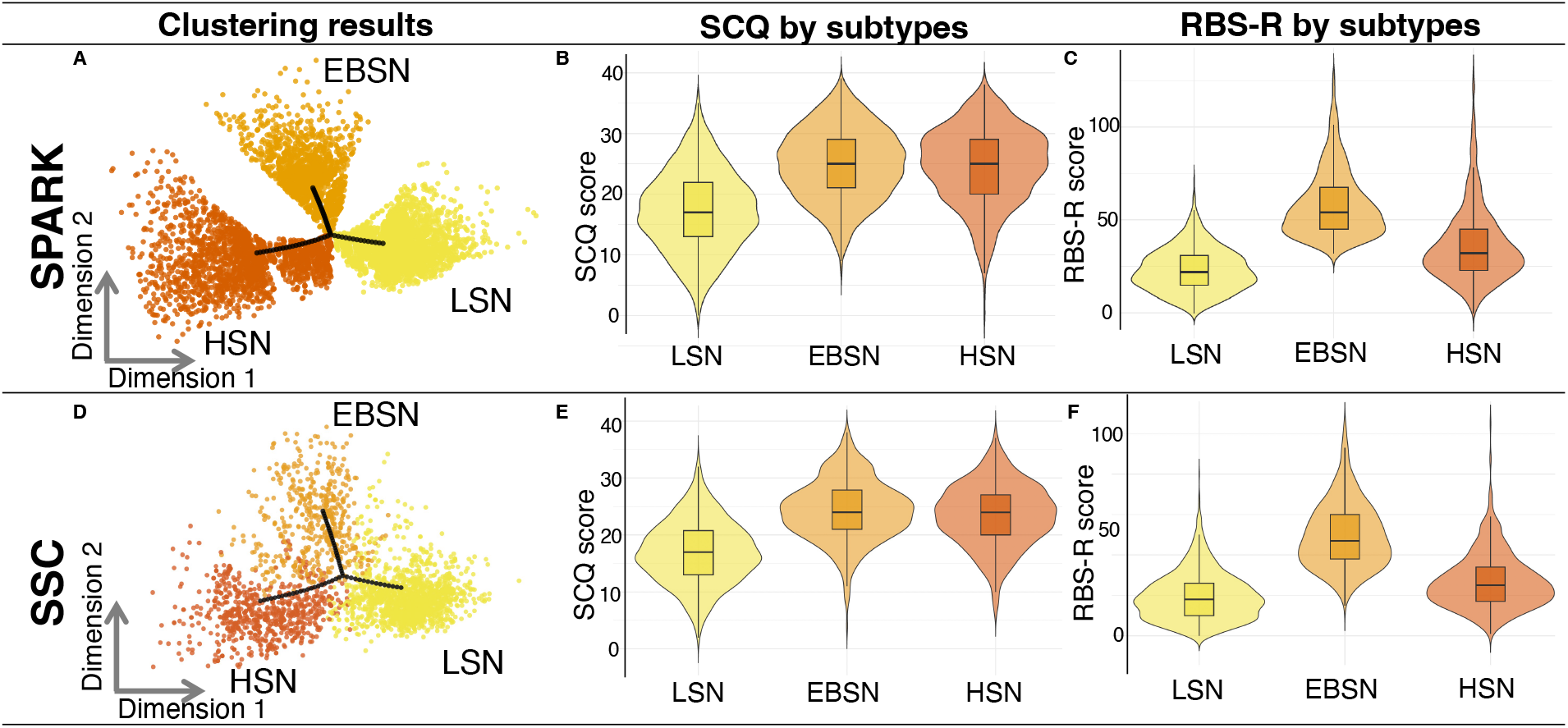
Projection of ML-derived ASD subtypes from SPARK to SSC. Panels A-C show the ML-derived ASD subtypes and the distribution of ASD core symptom scores across subtypes in SPARK, whereas Panels D-F show the corresponding results in SSC. Panels A and D show the distribution of autistic individuals assigned to the three ML-derived ASD subtypes (HSN, EBSN, and LSN) in the SPARK and SSC cohorts, respectively. Panels B and E show the distribution of Social Communication Questionnaire (SCQ) scores across ASD subtypes in the SPARK and SSC cohorts. Panels C and F show the distribution of Repetitive Behavior Scale-Revised (RBS-R) scores across the ML-derived ASD subtypes in the SPARK and SSC cohorts. Violin plots represent score distributions, with embedded boxplots indicating the median and interquartile range.

The projected subtype scores in SSC also exhibited similar patterns of phenotypic associations as SPARK (**Table 3**). HSN scores were positively associated with social communication deficits (r = 0.45, *P* < 2 × 10^-16^) and weakly with repetitive behaviors (r = 0.11, *P* = 5.43 × 10^-9^), while showing strong negative correlations with language ability (r = −0.57, *P* < 2 × 10^-16^) and cognitive performance (r = −0.69, *P* < 2 × 10^-16^; **Table 3**). Similarly, EBSN scores were strongly associated with repetitive behavior scores (r = 0.58, *P* < 2 × 10^-16^) and moderately with social communication deficits (r = 0.27, *P* < 2 × 10^-16^), showed minimal correlation with cognitive performance (r = 0.10, *P* = 6.84 × 10^-8^), and were associated with higher ADHD (r = 0.24, *P* < 2 × 10^-16^) and mood or anxiety symptom scores (r = 0.27, *P* < 2 × 10^-16^; **Table 3**). Conversely, LSN scores showed positive associations with language ability, cognitive performance, motor coordination, and adaptive functioning, together with negative associations with social communication deficits, repetitive behaviors, ADHD and mood disorders (**Table 3**). Overall, these continuous subtype score profiles closely mirrored those observed in SPARK, supporting the cross-cohort generalizability of the subtype framework.

Medication use in SSC, which was not included for subtype projection, also exhibited subtype-specific patterns similar to those observed in SPARK. Higher EBSN scores were associated with greater use of antidepressants (OR = 1.18, *P* = 9.67 × 10^-4^), and were also associated with medications for attention deficit disorders use (OR = 1.21, *P* = 1.83 × 10^-6^; **Table 4**). Higher HSN scores were strongly associated with sedative use (OR = 1.31, *P* = 5.22 × 10^-5^) and mood stabilizer use (OR = 1.28, *P* = 2.48 × 10^-9^; **Table 4**), whereas associations with antidepressants and antihypertensive medications were limited. In contrast, higher LSN scores were consistently associated with reduced medication use, including antidepressant (OR = 0.86, *P* = 1.89 × 10^-4^) as well as mood stabilizers (OR = 0.71, *P* = 5.66 × 10^-16^).

Together, these findings demonstrate that the ML-derived ASD subtype framework is transferable across independent cohorts despite differences in cohort composition and ascertainment. The projected subtype assignments and continuous subtype scores recapitulated the phenotypic and treatment-related characteristics observed in SPARK, providing a robust foundation for subsequent genetic analyses in both SPARK and SSC.

### 3.6. SNP heritability differs by ML-derived ASD subtype scores

We next investigated the genetic architecture of these ML-derived ASD subtypes among common and rare variants. SNP heritability was highest in the EBSN score (**h²g = 0.224, SE = 0.051**), followed by the HSN score (**h²g = 0.167, SE = 0.049**), which were higher than SNP heritability of 0.118 previously reported for the binary ASD diagnosis[57]. The LSN score showed a close-to-zero SNP heritability (**h²g = 0.064, SE = 0.044**). Due to limited sample size in SSC, heritability estimates for ASD subtype scores had large standard errors and were therefore not interpretable.

Additionally, we conducted the heritability enrichment analyses for HSN and EBSN scores using 927 cell-type-specific annotations from regulatory regions from 30 fetal and adult tissues including brain derived from ChIP-seq and ATAC-seq data in ENCODE and single-cell atlases[44]. We hypothesized that these two ASD subtype scores show different patterns of heritability enrichment across cell types, and we found that SNPs associated with the HSN score were enriched in regulatory regions of fetal excitatory neurons, fetal astrocytes and an adult prostate cancer cell line, whereas EBSN score associated SNPs were enriched in regulatory regions of fetal endodermal cells and adult motor neurons (*P* <0.05; **Table S4**).

Together, these findings suggest that the ML-derived ASD subtype scores capture distinct genetic architectures, with differences in both SNP heritability and heritability enrichment in specific cell types. These findings warrant further validation in cohorts with larger sample sizes.

### 3.7. Polygenic scores differ across ML-derived ASD subtypes

To account for potential confounding, we assessed the associations between ML-derived ASD subtype scores and PGS with adjustment for sex, age, and genetic ancestry using linear regression (**Table S5, Figure 6**). ADHD PGS was positively associated with increased EBSN scores in both SPARK (β = 0.08, *P* = 6.39 × 10^-8^) and SSC (β = 0.03, *P* = 3.52 × 10^-2^; **Table S5**). Likewise, Major

**Figure 6.**
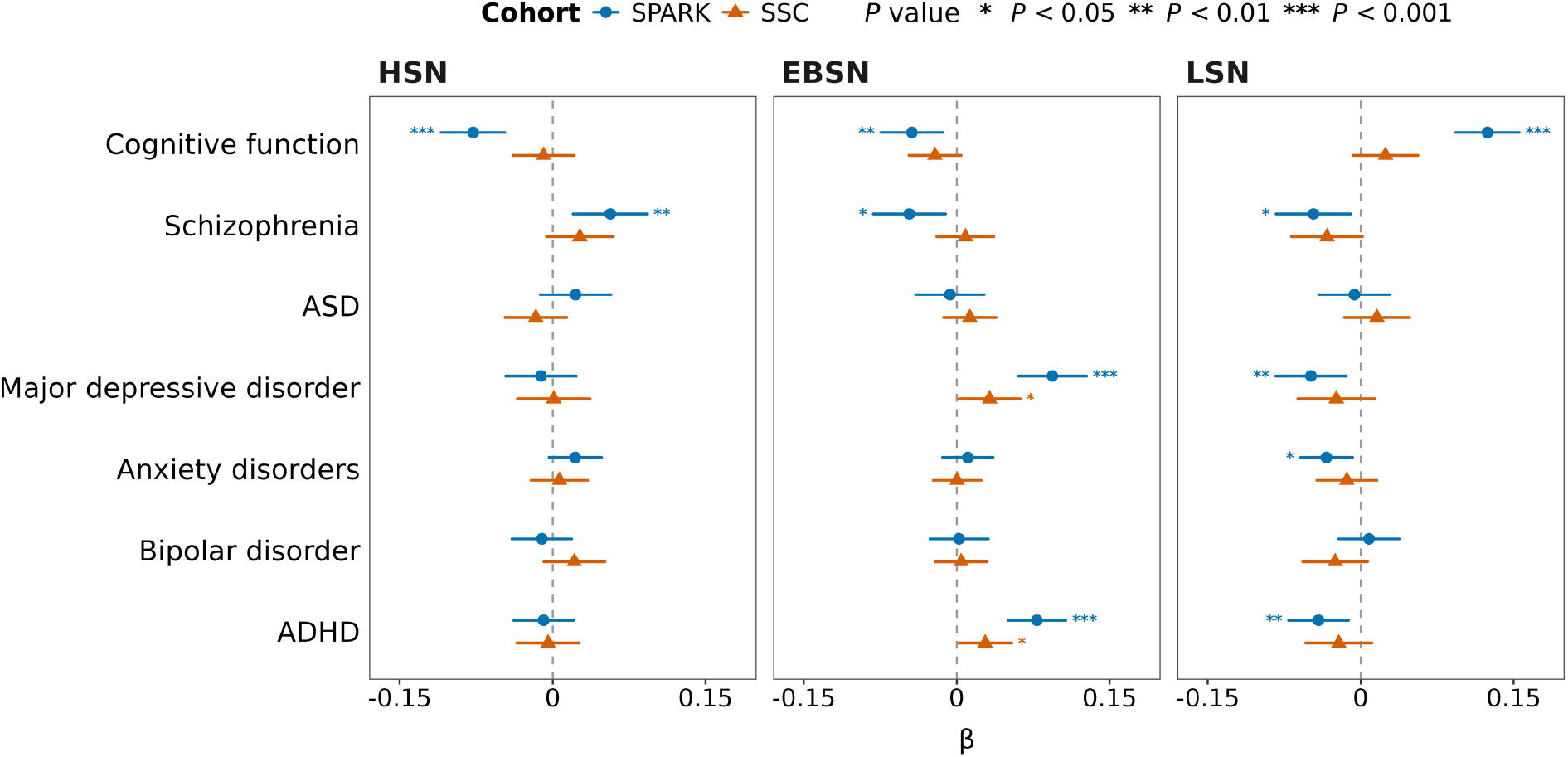
Associations between polygenic scores and continuous ASD support subtype scores in the SPARK discovery and SSC replication cohorts. The three panels show associations of standardized polygenic scores (PGS) for cognitive function, schizophrenia, autism spectrum disorder (ASD), major depressive disorder (MDD), anxiety disorders, bipolar disorder, and attention-deficit/hyperactivity disorder (ADHD) with the continuous LSN, EBSN, and HSN scores, respectively. Associations were evaluated separately in SPARK and SSC using linear regression models with ML-derived ASD subtype scores as the outcomes, adjusting for sex, age, and the first ten ancestry principal components(PCs). Points represent regression coefficients (β), and horizontal error bars indicate 95% confidence intervals. Blue circles represent SPARK estimates, and orange triangles represent SSC estimates. Positive β values indicate positive associations with the corresponding subtype score, whereas negative β values indicate inverse associations. The vertical dashed line indicates β = 0. Asterisks denote nominal statistical significance: *P < 0.05, **P < 0.01, and ***P < 0.001.

Depressive Disorder (MDD) PGS showed positive associations with EBSN scores across both cohorts (SPARK: β = 0.09, SE = 0.02, *P* = 4.25 × 10^-8^; SSC: β = 0.03, SE = 0.02, *P* = 3.65 × 10^-2^; **Table S5**). In SPARK, schizophrenia PGS was positively associated with the HSN score (β = 0.06, SE = 0.02, *P* = 2.33 × 10^-3^; **Table S5**), and PGS of cognitive function was negatively associated with both HSN and EBSN scores (both *P* < 10^-2^). In contrast, LSN scores were negatively associated with PGS for schizophrenia, ADHD, MDD and anxiety (all *P* < 0.05; **Table S5**). Similar directional associations were observed in SSC, although they did not reach statistical significance (**Table S5**).

Together, these findings demonstrate that the ML-derived ASD subtypes capture distinct polygenic architectures that are generally reproducible between SPARK and SSC. The consistent subtype-specific associations with neuropsychiatric PGS suggest that the ML-derived ASD subtype framework captures biologically meaningful dimensions of ASD heterogeneity.

### 3.8. Rare CNV burden is associated with HSN and EBSN subtypes

Rare exonic CNVs were called from WES data in SPARK [37, 38], and rare CNVs were called from WGS data in SSC as previously described in [39]. In SPARK, the mean exonic CNV burden is 7.42 per sample, with an average of 3.08 deletions and 4.34 duplications per sample. In SSC, the mean genome-wide CNV burden was 14.77 per sample, with an average of 12.07 deletions and 2.70 duplications per sample.

In SPARK, autistic individuals carrying one additional exonic CNV were associated with 4.2% higher odds of belonging to the HSN subtype compared with the LSN subtype (OR = 1.042, 95% CI 1.012–1.072; *P* = 0.006; **Table 5**) after adjusting sex, age and ancestry PCs. A similar association was observed for rare exonic duplications, with each additional exonic duplication associated with 5.8% higher odds of HSN versus LSN membership (OR = 1.058, 95% CI: 1.014– 1.103; *P* = 0.009; **Table 5**). The scores-based analyses were concordant with this pattern. Greater total exonic CNV burden (β = 0.003, 95% CI 0.001–0.006; *P* = 0.013) and greater exonic duplication burden (β = 0.004, 95% CI 0.001–0.008; *P* = 0.013; **Table 5**) were each nominally associated with a higher HSN score after adjusting sex, age and ancestry PCs.

In SSC, greater total CNV burden was associated with 2.6% higher odds of EBSN versus LSN membership (OR = 1.026, *P* = 0.001) and 3.0% lower odds of HSN versus EBSN membership (OR = 0.970, *P* < 0.001). Greater deletion burden was also associated with lower odds of HSN versus EBSN membership (OR = 0.972, *P* = 0.003). Consistent with these pairwise results, greater total CNV and deletion burdens were associated with higher EBSN scores (both β = 0.002, *P* < 0.05; **Table 5**)

### 3.9. *De novo* mutation burden is associated with high-support-needs subtypes in SPARK

We assessed the *de novo* mutation burden across ML-derived ASD subtypes in SPARK and SSC (**Table 5**). In SPARK, autistic individuals in the HSN subtype exhibited a significantly greater overall *de novo* variant (DNV) burden than those in the LSN subtype (OR = 1.10, *P* = 0.033). As for subtype scores in SPARK, greater overall DNV burden was associated with higher HSN scores (β = 0.040, *P* = 0.032). No significant subtype differences were observed for loss-of-function (LoF) DNV burden. In SSC, neither overall DNV burden nor LoF DNV differed significantly across ML-derived ASD subtypes (**Table 5**). Together, these findings suggest that the DNV burden among HSN subtypes is higher than other ASD subtypes.

## 4. Discussion

In this study, we developed a transferable ML-derived ASD subtype framework, moving beyond the diagnostic category to identify phenotypic dimensions that link clinical presentation to underlying genetic architecture. We developed an effective subtyping model that captures the multi-dimensional heterogeneity of symptoms among autistic children through three distinct ASD subtypes, as well as novel continuous subtype scores that will enable quantitative and genomic analyses. These ML-derived subtypes were associated with distinct support needs, patterns of intervention and special education service use, highlighting the validity and potential clinical utility of both the subtype classifications and scores. Importantly, we successfully projected this ASD subtype framework onto an independent cohort, where the projected subtype classifications and scores preserved the distinct clinical characteristics of the subtypes and consistently captured subtype differences in medication use. Our common and rare variant analyses among ∼5,000 subjects provided preliminary evidence of differences in the underlying genetic architecture across these ML-derived ASD subtypes, warranting further validation in larger cohorts.

The three ML-derived ASD subtypes reflected distinct clinical characteristics, rather than ordinal levels along a singular severity dimension. The HSN subtype was characterized predominantly by greater language impairment, lower cognitive performance, and reduced adaptive functioning, while the LSN subtype captured the opposite pattern. In contrast, the EBSN subtype was uniquely distinguished by more severe repetitive behaviors and co-occurring psychiatric comorbidities. This finding echoes previous subtypes found by Litman et al. [15], in emphasizing the often overlooked but critical role of mental health vulnerability in meaningful, tractable individual differences on the spectrum.

We were able to show continuous patterns of phenotypic variation within each subtype, consistent with prior evidence [8,9], and our framework captures this complexity by constructing continuous ASD subtype scores. While categorical subtypes facilitate clinical interpretation, continuous subtype scores preserve within-subtype variation and quantify the relative contribution of each subtype dimension for every individual. Compared with previous approaches that define ASD subtypes as discrete categories [8,15], these continuous scores preserve more individual-level variation and may provide greater power to detect genetic associations without partitioning the sample into distinct groups.

Another strength of our approach is that we developed a novel method to transfer the ML-derived ASD subtype framework by DDRTree across independent cohorts. The projected subtypes preserve consistent clinical profiles and facilitate cross-cohort comparisons. Our approach only requires similar domain-level symptom measures to project ASD subtype classifications and scores in an independent cohort, whereas previous approaches [8,15] required the same measures and item-level variables to reconstruct ASD subtypes in other cohorts. For example, in our study, although SSC did not include the same measurements on language and cognitive ability as SPARK, we successfully projected the framework onto SSC and identified subtypes with symptom profiles comparable to those observed in SPARK. This flexibility supports the application of our ASD subtype framework to larger and more diverse cohorts and provides a foundation for large-scale genetic studies of ASD heterogeneity.

These continuous subtype scores are promising for providing quantitative measures of support needs that complement the observations of parents, teachers and clinicians, judging by their clinical correlates demonstrated here. Although caregivers and educators often recognize the severity of a child’s challenges, these needs can be difficult to communicate objectively to other stakeholders. ASD subtype scores could help quantify individual support needs for the sake of eligibility, programming, and public health and advocacy.

Analyses of common and rare genetic variation provided preliminary evidence of distinct genetic architectures across the ML-derived ASD subtypes. The EBSN subtype score exhibited the highest SNP heritability and was associated with PGS of ADHD and MDD in both SPARK and SSC. Recent common variant analyses across 14 psychiatric disorders have identified a high genetic overlap between ASD and ADHD but relatively limited overlap between ASD and MDD [58]. Thus, autistic individuals with higher EBSN scores may carry a higher common variant burden. In contrast, although the HSN subtype also showed substantial SNP heritability, its overall genetic profile differed from that of EBSN. Consistent with its clinical profile of lower cognitive ability, more severe language impairment, and reduced adaptive functioning, the HSN subtype score was associated with lower GCF polygenic scores together with increased burden of both *de novo* variants and copy number variants, particularly duplication burden in SPARK. This pattern is consistent with previous studies that high-impact *de novo* and rare deleterious variants in autistic individuals are associated with cognitive impairment, developmental disabilities, and delayed developmental milestones [8, 9, 42].

Several limitations should be considered when interpreting these findings. First, the sample sizes for the genetic analyses were limited, which may have contributed to inconsistent results in SPARK and SSC. Some of the preliminary findings in SPARK were replicated in SSC, while others were nonsignificant or differed between the two cohorts. These discrepancies may reflect differences in inclusion and exclusion criteria in SPARK and SSC. SSC includes only simplex families and excludes autistic individuals with a low mental age, brain abnormalities, or birth defects [56], whereas SPARK includes both simplex and multiplex families, as well as autistic individuals with severe cognitive impairments. Moreover, the CNV results were not directly comparable between SPARK and SSC since CNVs were called from WES data in SPARK and WGS data in SSC. Therefore, these genetic findings should be interpreted cautiously and independently validated in larger cohorts. Second, the ML-derived subtypes were based on selected symptom measures that may not fully capture the full clinical and medical spectrum of ASD. Future studies could incorporate additional neurological and physical comorbidities such as sleep disorders and obesity, as well as other data modalities like neuroimaging, to provide a more comprehensive characterization of ASD heterogeneity. Finally, although symptom measures were standardized by sex, further analyses are needed to examine sex-specific patterns of symptom presentation.

Our study provides a transferable framework for characterizing ASD heterogeneity through clinically interpretable ML-derived subtype classifications and continuous subtype scores. This approach enables heterogeneous autistic populations to be examined within a unified subtype framework, which lays a good foundation for cross-cohort analyses and genetic studies of ASD heterogeneity [11,14]. Future studies identifying genetic risk factors associated with these ML-derived ASD subtypes may further inform personalized services and support as well as targeted early interventions for autistic individuals with heterogeneous clinical presentations.

## Data availability

The phenotypic and genetic data analyzed in this study were from the Simons Foundation Powering Autism Research for Knowledge (SPARK) study and the Simons Simplex Collection (SSC) through SFARI Base (https://base.sfari.org). Qualified researchers can request access to the SPARK and SSC population datasets used in this study through SFARI Base, subject to approval and applicable data use agreements.

## Code availability

The scripts for projecting ASD subtypes and subtype scores from one cohort to another is available on GitHub at https://github.com/AprilShuLab/DDRTreeProjection. The repository includes the computational methods and scripts used to reproduce the analyses and figures presented in this study, subject to access to the underlying datasets.

## Author contributions

Conceptualization, C.A.S., N.M., C.K.C., L.G.S.; methodology, Q.T., J.X., N.M., C.K.C., C.A.S., R.C., L.Y., F.C., B.T.; software, Q.T.; validation, Q.T., J.X.; formal analysis, Q.T., J.X.; investigation, Q.T., J.X.; resources, C.A.S.; data curation, C.A.S.; writing-original draft preparation, Q.T., J.X., C.A.S.; writing-review and editing, Q.T., J.X., L.G.S., R.C., L.Y., F.C., B.T., B.F., C.C., N.M., C.A.S.; visualization, Q.T.; supervision, C.A.S., N.M., C.K.C.; project administration, C.A.S.; funding acquisition, C.A.S. All authors reviewed and approved the final manuscript.

## Acknowledgements

We are grateful to all participating families and to the clinical sites and staff involved in the SPARK and SSC studies for their contributions to this research.

---

**Supplementary Figure 1.**
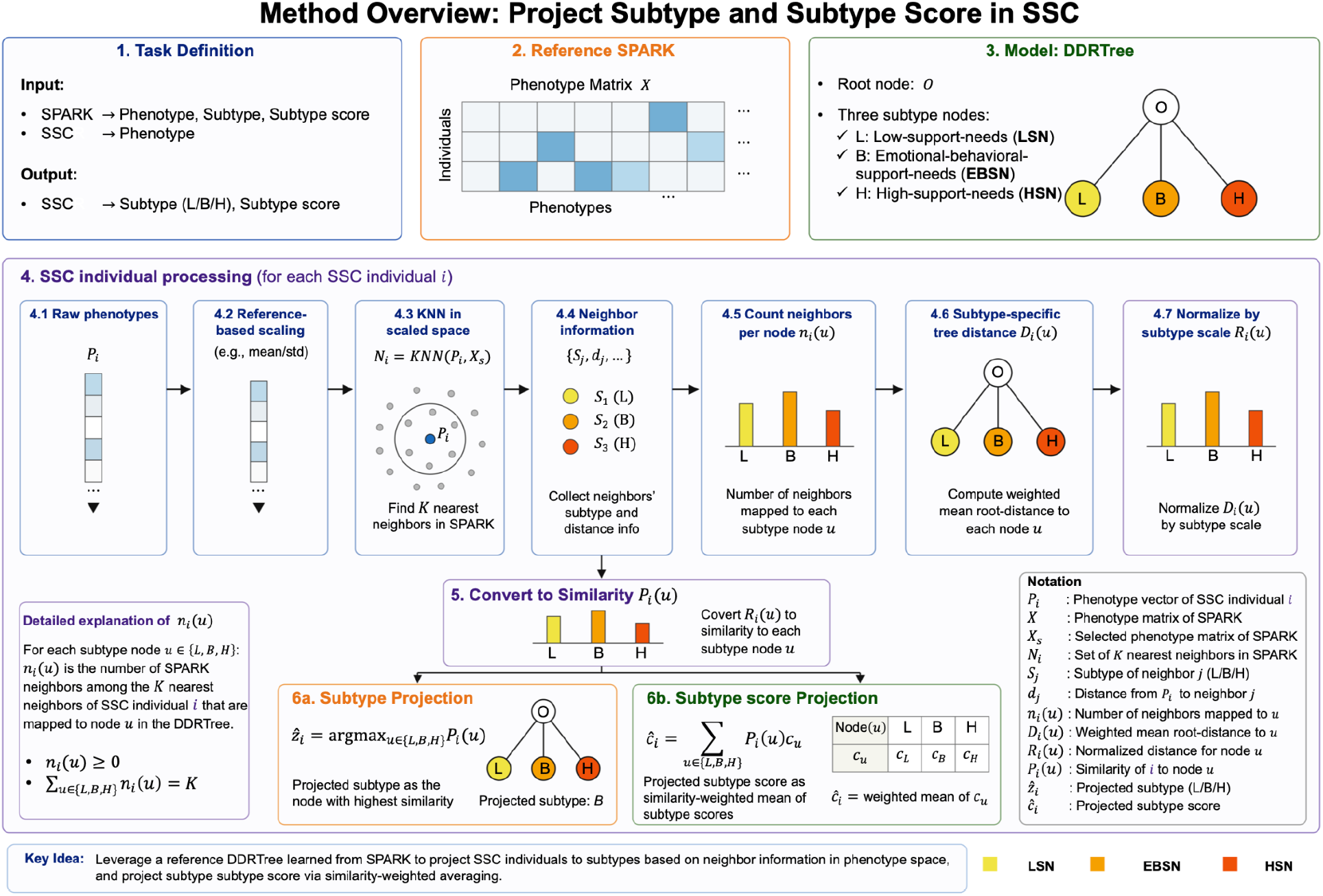
Tree-based KNN mapping framework for reference projection of ASD support-need subtypes in the SSC cohort. A reference DDRTree manifold was first constructed in the SPARK discovery cohort, defining three support-need subtype nodes (Low-support-need, Emotional-Behavioral-support-need, and High-support-need) together with corresponding continuous support-need scores. For each SSC participant, phenotypic measures were standardized using the scaling parameters estimated from the SPARK reference cohort before identifying the K nearest SPARK participants in the shared phenotype space. Neighboring reference individuals contributed both subtype membership and their positions within the reference DDRTree. Subtype-specific weighted root-to-node distances were calculated, normalized by branch-specific scales, and converted into similarity scores for each support-need subtype. The categorical subtype was assigned according to the highest similarity score, whereas continuous support-need scores were estimated as similarity-weighted averages of the corresponding SPARK subtype scores.

