## Supplementary Notes for "Machine Learning-Derived Subtypes of Autism Reveal Distinct Genetic Contributions from Common and Rare Variants"

### Supplementary Methods

#### 1. ML-derived ASD subtype classifications using DDRTree

To characterize heterogeneity in multidimensional ASD phenotypes, we applied discriminative dimensionality reduction via learning a tree (DDRTree [1]), as implemented in Monocle 2 [2]. DDRTree is a reversed graph embedding approach that jointly learns a low-dimensional representation of high-dimensional observations and a principal tree capturing their underlying structure. Its optimization objective is defined as follows:

$$\min_{\boldsymbol{W},\boldsymbol{B},\boldsymbol{R},\boldsymbol{Y},\boldsymbol{Z}} \sum_{i=1}^{N} \|x_i - \boldsymbol{W} z_i\|_2^2 + \frac{\lambda}{2} \sum_{k,k'} b_{k,k'} \|W y_k - W y_{k'}\|_2^2 + \gamma \left[ \sum_{k=1}^{K} \sum_{i=1}^{N} r_{i,k} \left( \|z_i - y_k\|_2^2 + \sigma \log r_{i,k} \right) \right] \quad (1)$$

Subject to $\boldsymbol{B}$ defines a spanning tree on the $K$ centroids

$$\boldsymbol{W}^{\boldsymbol{T}} \boldsymbol{W} = I_d$$

$$\sum_{k=1}^{K} r_{i,k} = 1 \text{ for each } i, r_{i,k} \geq 0, \forall i, k$$

where $X = \{\boldsymbol{x}_1, \dots, \boldsymbol{x}_n\} \in \mathbb{R}^{D \times N}$ contains the phenotypic profiles of $N$ individuals across d features after adjustment for se and age. The matrix $\boldsymbol{Z} = [\boldsymbol{z}_1, \dots, \boldsymbol{z}_N] \in \mathbb{R}^{d \times N}$ contains their corresponding representations in a $d$-dimensional latent space, and $\boldsymbol{W} \in \mathbb{R}^{D \times d}$ maps latent coordinates to the original feature space. The matrix $\boldsymbol{Y} = [\boldsymbol{y}_1, \dots, \boldsymbol{y}_K] \in \mathbb{R}^{d \times K}$ contains the latent centroids that serve as nodes of the principal tree. The matrix $\boldsymbol{B} = \{b_{kk'}\}$ specifies the tree edges connecting these centroids, while $\boldsymbol{R} = \{r_{ik}\}$ contains the soft-assignment weights linking individuals to centroids.

We applied this framework to phenotypic profiles of individuals in the SPARK cohort. The resulting principal tree summarized the major patterns of phenotypic variation across individuals, with distinct regions and branches corresponding to different multidimensional phenotypic profiles. Three major branches were identified and used to define the ML-derived ASD subtypes. Based on their clinical profiles and patterns of support needs, as described in the main text, these subtypes were designated as low-support-needs (LSN), emotional-behavioral-support (BSN), and high-support-needs (HSN).

#### 2. ML-derived ASD subtype scores

To complement the categorical subtype assignments, we derived continuous ASD subtype scores using the pseudotime framework implemented in Monocle 2. Following DDRTree reconstruction, individuals were projected onto the learned principal tree, and pseudotime was calculated as the cumulative distance along the tree from a specified root. As defined by Qiu et al. [23], pseudotime is calculated recursively as follows:

$$\phi_t(b_x, s_i) = \begin{cases} 0, & b_x = 1, i = 0 \\ \phi_t(Parent(b_x, s_i)) + \left\| \vec{p}(b_x, s_i), \vec{p}(Parent(b_x, s_i)) \right\|_2^2 & i > 0 \end{cases} \quad (2)$$

Where $\phi_t(b_x, s_i)$ denotes the pseudotime of individual $i$ along the corresponding trajectory segment, $Parent(b_x, s_i)$ denotes its parent on the minimum spanning tree of projected points, and $\vec{p}(b_x, s_i)$ represents its projected position on the trajectory. Pseudotime therefore measures distance along the tree rather than direct Euclidean distance in the original phenotypic space. In this application, it describes relative phenotypic position and doesn't imply temporal ordering or developmental progression.

We used this tree-distance framework to derive a continuous score for each of the three ASD subtypes. For each subtype, we identified the individual at the corresponding phenotypic extreme of the principal tree and calculated pseudotime for all individuals using the $orderCells()$ function in Monocle 2. The values were oriented so that the subtype-specific extreme corresponded to the maximum pseudotime, with larger values indicating positions closer to that extreme. The resulting values were rescaled to the interval [0,1] to obtain the corresponding ASD subtype scores. This procedure was repeated for the LSN, BSN, and HSN extremes, yielding three scores for each individual. These scores complemented the subtype assignments by quantifying continuous variation in phenotypic position relative to each subtype-associated phenotypic extreme.

#### 3. Projection of ML-derived ASD subtype classifications and ASD subtype scores from SPARK to SSC

To evaluate the reliability of the phenotypic structure identified in SPARK in an independent cohort, we developed a reference-based mapping framework to project both subtype assignments and ASD subtype scores from SPARK to SSC. The DDRTree structure, subtype branch definitions and individual ASD subtype scores derived from SPARK was retained as a fixed reference, while SSC participants were treated as query samples and didn't contribute to reconstruction of the reference trajectory. For each SSC participant, we first identified neighboring SPARK participants in the standardized phenotypic space. Their positions on the reference tree were then used to calculate similarity scores for the LSN, BSN, and HSN branches and determine the SSC participant's subtype assignment. Continuous ASD subtype scores were estimated through similarity-weighted aggregation of the corresponding scores of neighboring SPARK participants. The mapping workflow is illustrated in Supplementary Fig. S1. The individual steps are described in detail as follows. (**Supplementary Figure 1**)

##### 3.1. Reference-based standardization of phenotypic measures

To place SPARK and SSC participants in a common phenotypic space, we standardized phenotypic measures in both cohorts using the means and standard deviations calculated from the SPARK reference cohort. Let $X_{jm}$ denote phenotype $m$ for SPARK participant $j$, and $P_{im}$ denote the corresponding value for SSC individual $i$. The standardized values were calculated as:

$$X_{jm}^{(s)} = \frac{X_{jm} - \mu_m^{\text{SPARK}}}{\sigma_m^{\text{SPARK}}} \tag{3}$$

$$P_{im}^{(s)} = \frac{P_{im} - \mu_m^{\text{SPARK}}}{\sigma_m^{\text{SPARK}}} \tag{4}$$

where $\mu_m^{\text{SPARK}}$ and $\sigma_m^{\text{SPARK}}$ denote the mean and standard deviation of phenotype $m$ in SPARK, respectively. Applying the same reference-derived transformation to both cohorts ensured that subsequent distance calculations used a common scale for each phenotypic measure.

##### 3.2. Identification of neighboring SPARK participants

For each SSC participant, we identified the *K* nearest SPARK participants using Euclidean distance in the standardized phenotypic space. The distance between SSC participant *i* and SPARK participant *j* was calculated as:

$$d_{ij} = \left\| P_i^{(s)} - X_j^{(s)} \right\|_2 \tag{5}$$

where $P_i^{(s)}$ and $X_j^{(s)}$ denote their respective standardized phenotypic vectors. Let $j_{i,1}, \dots, j_{i,N}$ denote the indices of SPARK participants ordered by increasing distance to SSC participant $i$, such that $d_{i,j_{i,1}} \leq \cdots \leq d_{i,j_{i,N}}$. The neighbor set was defined as:

$$\mathcal{N}_i = \{j_{i,1}, \dots, j_{i,K}\} \tag{6}$$

Thus, $\mathcal{N}_i$ contains the indices of the *K* SPARK participants closest to SSC participant $i$ in the standardized phenotypic space. Each neighbor's subtype assignment and projected position on the reference tree were retained for subsequent mapping.

##### 3.3. Aggregation of neighboring SPARK participants by subtype

For each SSC participant, the *K* neighboring SPARK participants were subsequently grouped according to their reference subtype assignments. Let

$$\mathcal{U} = \{L, B, H\} \tag{7}$$

denote the labels corresponding to the LSN, BSN, and HSN subtypes, respectively. For SSC individual *i*, the number of neighboring SPARK participants assigned to subtype *u* was calculated as:

$$n_i(u) = \sum_{j \in N_i} \mathbf{1}\{z_j = u\}, \qquad u \in \mathcal{U} \tag{8}$$

Where $z_j$ is the subtype assignment for SPARK participant *j*, and $\mathbf{1}\{\cdot\}$ is the indicator function. Because each neighbor belonged to exactly one subtype, we have

$$\sum_{u \in \mathcal{U}} n_i(u) = K. \tag{9}$$

This step characterized the subtype composition of each SSC participant's reference neighborhood. The neighbors' positions on the reference tree were then used to calculate subtype-specific tree distance, as described below.

##### 3.4. Calculation of subtype-specific DDRTree branch distances

For each SSC participant, we summarized the positions of neighboring SPARK participants on each subtype branch using a weighted mean distance from the common root. Let $\mathfrak{B}_u$ denote the set of reference tree nodes belonging to subtype branch $u \in \mathcal{U}$ and let $Q_i(u) \subseteq \mathfrak{B}_u$ contain the nodes to which at least one neighbor of SSC participant *i* was projected. If $c_{iq}$ denotes the number of neighbors projected to node *q*, the total number of neighbors on branch *u* is

$$n_i(u) = \sum_{q \in Q_i(u)} c_{iq}. \tag{10}$$

Let $\boldsymbol{Y_q}$ and $\boldsymbol{Y_O}$ denote the latent coordinates of node *q* and the common root *O*, respectively. The root-to-node distance was calculated as

$$d_q^{tree} = \left\| \boldsymbol{Y_q} - \boldsymbol{Y_O} \right\|_2. \tag{11}$$

Nodes within the branch were weighted by the proportion of neighbors on that branch projected to it:

$$w_{iq}(u) = \frac{c_{iq}}{n_i(u)}, q \in Q_i(u), \tag{12}$$

such that

$$\sum_{q \in Q_i(u)} w_{iq}(u) = 1. \tag{13}$$

The subtype-specific branch distance was then defined as the weighted mean root-to-node distance across the occupied nodes on branch $u$:

$$D_i(u) = \sum_{q \in Q_i(u)} w_{iq}(u)\, d_q^{tree} \tag{14}$$

If none of the $K$ nearest SPARK neighbors was projected to a node belonging to subtype branch $u$, $D_i(u)$ was set to zero. Thus, $D_i(u)$ summarizes how the neighboring SPARK participants are distributed along subtype branch $u$ relative to the common DDRTree root, rather than representing the distance to any single DDRTree node. Larger values indicate that these neighbors lie farther from the common root in the latent space.

##### 3.5. Normalization and derivation of subtype similarity scores

To account for differences in the geometric scales of the three subtype branches, we normalized each branch distance by a reference branch scale. For subtype $u$, the branch scale $s_u$ was defined as the distance from the common root $O$ to the most distal node on that subtype branch:

$$s_u = \left\| \boldsymbol{Y}_{\boldsymbol{q}_u^*} - \boldsymbol{Y}_{\boldsymbol{O}} \right\|_2 \tag{15}$$

where $\boldsymbol{q}_u^*$ denotes the most distal node on subtype branch $u$. For SSC individual $i$, the normalized branch distance was then calculated as:

$$R_i(u) = \frac{D_i(u)}{s_u}, u \in \mathcal{U} \tag{16}$$

Thus, $R_i(u)$ represents the relative progression of the subtype-specific neighboring SPARK participants along branch $u$ after accounting for differences in branch scale. The normalized branch distances were subsequently rescaled across the three subtype branches to obtain subtype-specific similarity scores:

$$P_i(u) = \frac{R_i(u)}{\sum_{v \in \mathcal{U}} R_i(v)}, u \in \mathcal{U} \tag{17}$$

The resulting subtype-similarity vector was defined as

$$\boldsymbol{P_i} = (P_i(L), P_i(B), P_i(H)) \tag{18}$$

Because $R_i(u)$ quantifies relative progression from the common root along subtype branch $u$, a larger $P_i(u)$ indicates greater similarity of SSC individual $i$ to the corresponding SPARK-derived subtype.

##### 3.6. Projection of ASD subtype classifications

Each SSC participant was assigned to the subtype with the highest similarity score:

$$\hat{z}_i = \underset{u \in \mathcal{U}}{\operatorname{argmax}} P_i(u) \tag{19}$$

Thus, subtype prediction was based on the relative DDRTree-informed similarity to the three subtype branches rather than on a simple majority vote among the $K$ nearest SPARK neighbors. Neighboring SPARK participants contributed to the final assignment through their projected positions within the reference tree, as incorporated in the subtype-specific branch distances and similarity scores described above.

##### 3.7. Projection of ASD subtype scores

For each SSC participant, the three continuous ASD subtype scores were estimated by weighted averaging of the corresponding scores of all $K$ neighboring SPARK participants. Each neighbor $j$ was first assigned the similarity score associated with its reference subtype label $z_j$:

$$a_{ij} = P_i(z_j) \tag{20}$$

These values were then normalized across the neighbor set to obtain:

$$W_{ij} = \frac{P_i(z_j)}{\sum_{l \in N_i} P_i(z_l)}, \tag{21}$$

Such that $\sum_{j \in N_i} W_{ij} = 1$. This normalization converts the subtype-level similarities into individual neighbor weights, accounting for the number of neighbors belonging to each subtype.

Let $c_j^{(r)}$ denote the SPARK-derived score of the participant $j$ for subtype-score dimension $r \in \mathcal{U}$. The projected subtype score for SSC individual $i$ was calculated as:

$$\hat{c}_i^{(r)} = \sum_{j \in N_i} W_{ij} c_j^{(r)} \tag{22}$$

The same calculation can be expressed by grouping neighbors according to their subtype assignments. Define $N_i(u)$ and scaled sum

$$c_i^{(r)}(u) = \frac{\sum_{j \in N_i(u)} c_j^{(r)}}{\sum_{l \in N_i} P_i(z_l)}. \tag{23}$$

Equation (22) can then be written in the subtype-level form shown in Supplementary Fig. S1:

$$\hat{c}_i^{(r)} = \sum_{u \in \{L,B,H\}} P_i(u) c_i^{(r)}(u)\,. \tag{24}$$

Here, $c_i^{(r)}(u)$ is denoted as cu in Supplementary Fig. S1 for simplicity. The calculation was performed separately for the Low-support-need, Behavioral-support-need, and High-support-need score dimensions, generating three projected ASD subtype scores for each SSC participant.

##### 3.8. Finalization of the SPARK-to-SSC projection framework

For each SSC individual $i$, the reference-based mapping procedure produced one subtype assignment and three subtype scores corresponding to the Low-support-need, Behavioral-support-need, and High-support-need dimensions:

$$O_i = \left\{\hat{z}_i, \hat{c}_i^{(L)}, \hat{c}_i^{(B)}, \hat{c}_i^{(H)}\right\} \tag{25}$$

The subtype assignment was determined by the highest branch similarity score, while the continuous scores were estimated by weighted averaging of neighboring SPARK participants' reference scores. The SPARK-derived DDRTree structure, subtype branch definitions, branch-specific scaling factors, and reference ASD subtype scores were retained as fixed reference information throughout the mapping procedure. SSC participants were used only as predict samples and did not contribute to reconstruction or optimization of the reference tree. The resulting subtype assignments and ASD subtype scores were subsequently used for phenotypic and genetic validation analyses in SSC. The complete mapping framework is summarized in Supplementary Fig. S1.

#### 4. Assessment of reliability of projected subtype assignment and scores

To evaluate the reliability of the SPARK-to-SSC projection, we performed a stratified-bootstrap analysis to assess the reliability of projected subtype assignments and subtype scores under resampling of the SPARK reference cohort. Bootstrap resampling was performed separately within each of the three SPARK-derived subtypes, thereby preserving the original subtype composition of the reference cohort across bootstrap replicates. Projection reliability was evaluated for neighborhood sizes ($k$ = 2, 3, 4, and 5), with five stratified-bootstrap replicates performed for each value of $k$.

For each bootstrap replicate $b$, SPARK individuals were sampled with replacement independently within each subtype. Let $A_u$ denote the set of SPARK individuals assigned to subtype $u \in \mathcal{U}$. For each subtype, a bootstrap sample $A_u^{(b)}$ was generated by sampling with replacement from $A_u$, while maintaining the original subtype-specific sample size:

$$A_u^{(b)} \sim Bootstrap(A_u), \qquad \left|A_u^{(b)}\right| = |A_u|. \tag{26}$$

The three subtype-specific bootstrap samples were then combined to construct the bootstrap SPARK reference cohort for replicate *b*. Using this resampled reference cohort, the complete tree-based k-nearest-neighbor projection procedure described in Section S1.3 was repeated to obtain a projected subtype assignment and three ASD subtype scores for each SSC individual.

The reliability of projected subtype assignments was evaluated based on the consistency of subtype labels across the five bootstrap replicates for each neighborhood size *k*. For SSC individual $i$, let $\hat{z}_i^{(b,k)}$ denote the subtype assignment obtained from bootstrap replicate *b* at neighborhood size *k*. The maximum number of replicates assigning individual *i* to the same subtype was defined as

$$C_i(k) = \max_{u \in \mathcal{U}} \sum_{b=1}^{5} I\left(\hat{z}_i^{(b,k)} = u\right). \tag{27}$$

Individuals with identical subtype assignments across all five bootstrap replicates, corresponding to $C_i(k) = 5$, were classified as having **high reliability**. Individuals assigned to the same subtype in four of five replicates, corresponding to $C_i(k) = 4$, were classified as having **moderate reliability**. Individuals for whom the most frequently assigned subtype occurred in three or fewer replicates, corresponding to $C_i(k) \leq 3$, were classified as having **low reliability**.

The reliability of projected ASD subtype scores was evaluated separately for each subtype score and neighborhood size *k*. For SSC individual $i$ and subtype *g*, let $\hat{s}_{iu}^{(b,k)}$ denote the projected subtype score obtained from bootstrap replicate *b* at neighborhood size *k*. The mean score across the five bootstrap replicates was calculated as

$$\bar{s}_{iu}^{(b,k)} = \frac{1}{5}\sum_{b=1}^{5} \hat{s}_{iu}^{(b,k)}. \tag{28}$$

The bootstrap standard deviation for each individual was then calculated as

$$SD_{iu}(k) = \sqrt{\frac{1}{4}\sum_{b=1}^{5}\left[\hat{s}_{iu}^{(b,k)} - \bar{s}_{iu}^{(b,k)}\right]^2}. \tag{29}$$

Smaller bootstrap standard deviations indicate greater reliability of the projected ASD subtype score for an individual. For each subtype score and neighborhood size *k*, the individual-level bootstrap standard deviations were further summarized across all SSC individuals by their mean:

$$SD_u(k) = \frac{1}{N_{SSC}}\sum_{i=1}^{N_{SSC}} SD_{iu}(k). \tag{30}$$

The resulting mean bootstrap standard deviation was used as the overall measure of subtype-score reliability, with smaller values indicating greater reliability across bootstrap-resampled SPARK reference cohorts.

Projection reliability was evaluated across neighborhood sizes *k* = 2~5 using the complementary measures defined above. Subtype reliability was summarized by the proportions of SSC individuals classified as having high, moderate, or low subtype-assignment reliability, whereas subtype-score reliability was summarized by the mean bootstrap standard deviation for each of the three subtype scores. These measures were jointly considered to compare projection robustness across different neighborhood sizes and to guide the selection of $k$ for the final SPARK-to-SSC projection.

### Figures

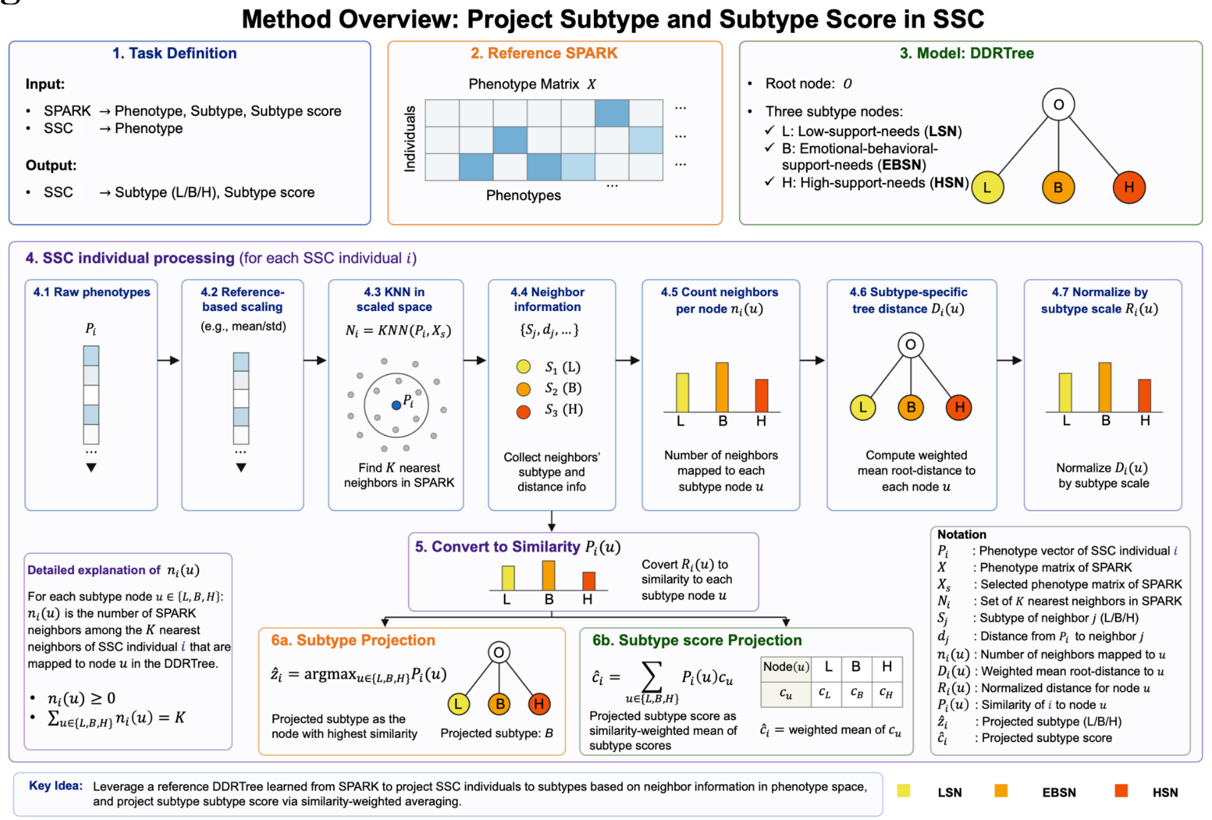

**Supplementary Figure 1. Tree-based KNN mapping framework for reference projection of ASD support-need subtypes in the SSC cohort.** A reference DDRTree manifold was first constructed in the SPARK discovery cohort, defining three support-need subtype nodes (Low-support-need, Emotional-behavioral-support-need, and High-support-need) together with corresponding continuous support-need scores. For each SSC participant, phenotypic measures were standardized using the scaling parameters estimated from the SPARK reference cohort before identifying the K nearest SPARK participants in the shared phenotype space. Neighboring reference individuals contributed both subtype membership and their positions within the reference DDRTree. Subtype-specific weighted root-to-node distances were calculated, normalized by branch-specific scales, and converted into similarity scores for each support-need subtype. The subtype was assigned according to the highest similarity score, whereas continuous support-need scores were estimated as similarity-weighted averages of the corresponding SPARK subtype scores.
